# Efficacy and Safety of Nebulized Ethanol Inhalation in COVID-19 Treatment. A Randomized, Clinical Trial

**DOI:** 10.1101/2022.06.15.22276427

**Authors:** Ali Amoushahi, Elham Moazam, Amin Reza Tabatabaei, Golnaz Ghasimi, Pietro Salvatori, Ian Grant-Whyte, Ahmed Ragab Ezz

**Author notes:** Corresponding Author; Department of Anesthesiology and Intensive Care Unit, Isabn-e-Maryam Hospital, Isfahan University of Medical Sciences, Isfahan, Iran.

## Abstract

**Background:** Considering anti coronavirus effects of ethanol, the efficacy of its administration was evaluated in this research. Because of respiratory tract entrance of virus in COVID-19, this study was done by inhalation of nebulized ethanol.

**Methods:** Ninety-nine positive SARS-CoV-2-PCR patients who had been admitted at a respiratory clinic were included in this study. Patients were randomly assigned to the control (distilled water spray) and intervention (35% ethanol spray) group. Both groups were instructed to inhale 3 puffs of spray and inhale it, every six hours for a week. Global symptomatic score (GSS), clinical status scale,0020Blood Oxygenation, and C-Reactive Protein (CRP) at the first visit and days 3, 7, 14 were measured and compared between groups.

**Results:** The GSS decreased more and faster in the intervention group (ethanol) (1.4+1.4 vs 2.3+1.7, P=0.035) two weeks after starting intervention. On day 14, the odds of intervention group to have better clinical status was 5.715 times (95% CI, 2.47 to 13.19) than of control group a statistically significant effect, Wald χ2 (1) =16.67, P =0.001. Blood oxygen saturation also improved earlier in the ethanol group but without statistical significance difference. The readmission rate was lower in the intervention group (zero vs 10.9%, P=0.02).

**Conclusion:** Inhaled ethanol seems to be effective in improvement, mitigating clinical symptoms and reducing the need to repeat treatment. Considering the low cost, availability and no significant adverse events of ethanol, research and additional efforts are recommended to evaluate its curative effects in the early stages of COVID-19.

## Background

The cytokine storm is the cause of many deaths in COVID -19. The antiviral effects of ethanol with solving the fat layer (1) and destroying the glycoprotein of coronavirus have already been established (2). Proven antiviral effects of ethyl alcohol on extracellular surfaces have been demonstrated by researchers (3). Immunological studies have shown that acute administration of ethanol can have immunomodulatory effects on innate immunity system mediated by TNFamRNA protein and mitogen-activated protein-kinase, together with lowered cytokine storm by reducing inflammatory factors such as TLR4, TLR8, TLR9, interleukin-6 and IL-3 (4, 5). Ethanol is also helpful with the chemotaxis of bronchoalveolar macrophages (6). Other demonstrated effects of ethanol are lowering virus replication by inhibition of RNA-dependent polymerase (7), bronchial dilation by relaxing involuntary smooth muscles (8), sedation and relaxation of the patient (9) and muscular analgesic effects (10).

Ethanol administration had previously been reported for the treatment of methanol poisoning (11), fat embolism (12), prevention of preterm labor (13), preeclampsia (14), and pulmonary edema (15). The histological safety of inhalation ethanol therapy in the lungs and respiratory tracts of rodents has been recently shown by Ana Castro-Balado et al (16). Ethanol is approved by the Food and Drug Administration. Given these effects of ethanol on virus wall destruction, inhibition of proliferation, and inhibition of immune hyperactivity, the question now is, “Can ethanol inhalation therapy be effective in controlling COVID-19?”

There is no prior knowledge of the inhalation ethanol therapy in COVID-19. This idea was first suggested and published one month after COVID-19 pandemic (17) in Iran (February 2020). Later, a paper dealing with the rationale of ethanol use in this field was presented (18). To try finding the answer, a clinical trial was conducted to evaluate the effectiveness of ethanol therapy on clinical status and prognosis in a defined set of patients. The study was approved by the Medical University of Isfahan, Research and Ethics Committees and was registered at https://irct.ir/trial/58201

## Methods

### Study Design and Oversight

This study is a randomized triple-blind clinical trial with a control group and a parallel design that was conducted at the Isabn-e-Maryam hospital of the Medical University of Isfahan, Iran, in September and October 2021. Patients were randomly assigned in a 1:1 ratio. The study was originally intended for patients admitted to the hospital, but due to amending the country’s policy of setting up respiratory clinics in hospitals and prescribing Remdesivir and dexamethasone to patients with moderate COVID-19, the study was conducted at this center in Isabn-e-Maryam Hospital. The protocol was approved by the Isfahan University of Medical Science Ethics Committee. (Code: IR.MUI.MED.REC.1400.506).

### Patients

The study population consisted of positive SARS-CoV-2 detected by RT-PCR test. They had moderate COVID-19 pneumonia and were referred to the Respiratory Clinic of the Hospital. Inclusion criteria were: agreeing to implement the plan of informed consent, age over 12 years, not pregnant, no history of asthma, alcoholism or epilepsy, no contraindications to ethanol.

Exclusion criteria were: intolerance to inhaled ethanol, hypersensitivity or allergies to ethanol, use of drugs that interact with ethanol, and partial or incomplete treatment. Ethanol patch skin test was used to detect possible allergy to alcohol. In this experiment, a drop of ethanol was placed on a gauze pad and taped it to the patient’s arm. After about seven minutes, signs such as redness, swelling or itching of the skin were sought. These signs indicated the possibility of allergy or intolerance to alcohol.

### Intervention

Both control and intervention groups were enrolled among patients addressed to the standard treatments indicated in the national clinical guidelines of Iran. The standard treatment was intramuscular dexamethasone, 8mg/day (5 days) and 200 mg of Remdesivir intravenously on day 1, followed by 100 mg of Remdesivir once daily for 4 subsequent days, infused over 30 to 60 minutes. Informed consent was obtained from all patients. In addition to the standard treatment distilled water spray (placebo) was added in the control group and 35% ethanol spray was added in the intervention group. Two sets of 100 ml spray were provided. All of them were instructed to spray three times every 6 hours from a distance of 20-30 cm from their face, while wearing a mask and closing their eyes, and to take a deep breath when they feel nebulized liquid droplets in their nose, mouth, throat, larynx and lungs. We emphasized them that this protocol must have been repeated daily, every six hours, up to 7 days. This procedure was first taught by nurses and, after health workers had verified the patient’s mastery, it was prescribed to the patient to be continued at home. Compliance with spray use was checked at every visit and patients who did not use it or used it irregularly were excluded from the study.

### Clinical and Laboratory Monitoring

The data collection sheet had two sections. In the first part, demographic and underlying diseases information were reported. In the second part, clinical information of research cases was recorded. Data collection checklists were completed by a trained nurse and included clinical symptoms, para-clinical results, clinical examinations and patient record contents. Data related to research variables including blood oxygen saturation by pulse oximetry, level of inflammatory factor (CRP), need for adjunctive treatment or readmission in hospital and clinical symptoms were collected for both groups until hospital discharge. The Global Symptomatic Score (GSS) was obtained by calculation of cumulative scores of clinical signs and symptoms including fever, headache, body aches, sore throat, nasal discharge, chills, cough, shortness of breath, anorexia, loss of smell and loss of taste. Any possible side effects were treated in both groups and reported if present. Patients’ oxygenation status was monitored and recorded daily with a pulse oximeter. The pulse oximeter was fixed, and at the time of measurement, the patient was breathing room air without receiving supplemental oxygen.

A modified 7-point ordinal scale was used to assess the clinical condition on day 14 of treatment period (19). This scale (CSS) has 7 indexes:

1. Death
2. Hospitalized, on invasive mechanical ventilation
3. Hospitalized, on non-invasive ventilation or high flow oxygen devices
4. Hospitalizations for any reason and need oxygen
5. Requiring ongoing medical care or supplemental oxygen at home
6. Continue signs or symptoms of COVID-19 without requiring supplemental oxygen - no longer requires ongoing medical care
7. Complete recovery

The need for hospitalization in the intensive care unit, drug side effects, clinical symptoms and mortality of the research samples were monitored in both groups. Side effects were recorded after the informed consent was signed and were graded based on version 5.0 of the Common Terminology Criteria for Adverse Event.

The final follow-up was performed on the 14th day of the disease through telephone calls and review of patients’ records and hospital information system documents. The outcome comparisons were performed at 0, 3, 7 and 14 days after the intervention.

### Sampling

Sampling was performed by an easy random method. Computerized Random number table was used for random assignment. One researcher determined the sequence of random allocations without coordination with others. This person was different from the nurses who assigned participants to the interventions. This person filled the sprays (nebulizer) one by one with 100ml of diluted distilled water or ethanol-35% and labeled them with the numbers coming out of the number container. She assigned a questionnaire and consent form for each spray and wrote the number on the spray in the questionnaire. Each spray was delivered to one of the participants in the study and his / her family or companion was instructed on how to use it. Blinding was performed at the level of patients, clinicians, care provider and analyst.

### Statistical Analysis

Based on the “treatment-on” or “protocol per” strategy, analysis was limited to participants who, according to the study protocol and inclusion criteria, received full interventions. Quantitative and qualitative variables were reported in the form of descriptive statistics including mean and standard deviation and number (%), respectively. Quantitative variables with normal and abnormal distributions were compared between groups using independent t-test and Mann-Whitney test, respectively. In addition, qualitative variables were compared between the two groups using Chi-square test.

## Results

### Patient Characteristics

In this study, 150 COVID-19 patients were evaluated for inclusion based on the positive result of RT-PCR test. Two patients did not meet the inclusion criteria and 24 patients did not agree with the study. One-hundred-twenty-four patients were divided into two groups (Intervention and Control). Twenty-five patients were excluded from the study: 6 patients due to intolerance to ethanol inhalation. Hiccups, eye irritation, cough, shortness of breath, sneezing and the unpleasant smell of alcohol were the main reasons for their intolerance. Nine-teen of these patients (10 in the intervention group and 9 in the control group) were excluded from the study due to irregularity or no compliance with the recommended method. Finally, a total of 99 patients were included in the study: 44 in the treatment group and 55 in the control group, respectively. Figure 1 shows the characteristics of the studied patients in the intervention and control groups.

**Figure 1.** Flowchart of the Study.

Ninety-nine patients of Covid-19 Respiratory Outpatient Clinic of Isabn-e-Maryam Hospital of Isfahan University of Medical Sciences were randomly assigned to intervention (44 patients) and control (55 patients) groups from September to November 2021. 56 patients (56.6%) were female. The mean age of patients was 46.4±12.8. Table 1 shows demographic and baseline characteristics of patients in the two groups.

**Table 1.** Demographic characteristics in two research groups.

| P value | Intervention group<br>N=44 | Control group<br>N=55 | Index |
| --- | --- | --- | --- |
| 0.928 | 45.91±12.58 | 46.15±13.15 | **Age (years) (Mean± SD)** |
| 0.804 | (20.5) 9<br>(50) 22<br>(22.7) 10<br>(6.8) 3 | 16 (29.1)<br>25 (45.5)<br>10 (20)<br>3 (5.5) | **BMI) Kg/m2)**<br>(Mean± SD)<br>Normal weight<br>Overweight<br>Obesity<br>Excessive obesity |
| 0.024 | ) 43.2( 19<br>(56.8) 25 | 37 )67.3 (<br>18 (32.7) | **Gender N (%)**<br>**Female**<br>**Male** |
| 0.251 | (16.3) 8<br>(14.3) 7<br>(32.7) 16<br>(30.6) 16<br>(6.1) 3 | (5.4) 3<br>(14.3) 8<br>(50) 28<br>(26.8) 16<br>(3.6) 2 | **Education Level N (%)**<br>Illiterate and<br>Elementary<br>Secondary<br>Diploma<br>Bachelor-higher<br>Unknown |
| 0.614 |  |  | **Risk factors for** |
|  | 38 )69.1 (<br>12 (21.8)<br>4 (7.3)<br>1 (1.8) | 23 )52.3 (<br>19 (43.2)<br>2 (4.5)<br>0 | **disease N (%)**<br>Not any<br>1 risk factor<br>2 risk factors<br>3 risk factors |

38 patients suffered from underlying diseases. The most common underlying disease in both groups was diabetes mellitus. 4 patients (7%) from the control group and 6 patients (14.3%) from the intervention group were diabetic patients. 7 patients had cardiovascular problems and 7 patients had hypertension.

### Clinical signs and symptoms at the time of admission

Patients also did not differ significantly in the time distance between the onset of symptoms and admission, pulmonary involvement, and early clinical signs and symptoms at baseline. The basic characteristics of patients’ clinical signs and symptoms are given in Table 2.

**Table 2.**
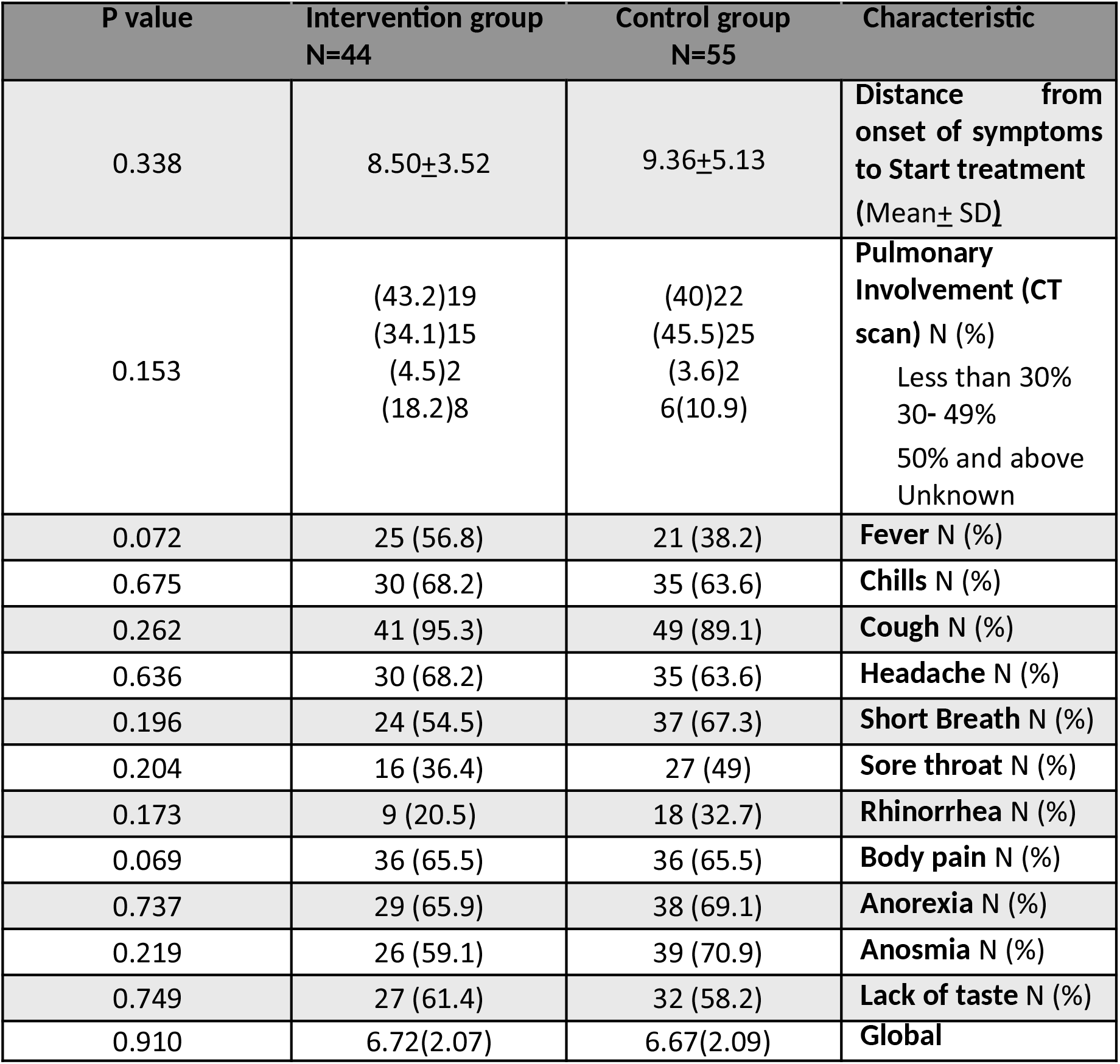
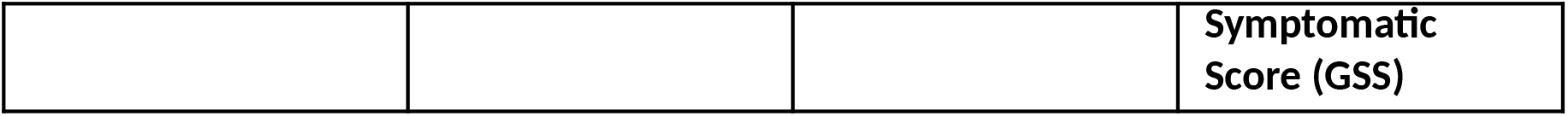
Preliminary characteristics of signs and symptoms, risk factors and laboratory values in baselines.

The most primary clinical symptoms in the intervention group were cough, body aches, chills and headache. Cough, olfactory disturbance and anorexia were more common in the control group. No significant difference was observed in symptoms.

### Evaluation of Global Symptomatic Score of the two groups after entering the research and intervention

Global Symptomatic Score (GSS) is considered as an indicator of symptoms and signs of the patients. GSS was assessed in both groups, at the beginning of treatment and after 3, 7, 14 days of treatment. The results are shown in Figure 2. Statistical analysis showed that the clinical status of the two groups was the same at the beginning of the study, but in the intervention group (ethanol therapy) the clinical symptoms decreased rapidly more than in the control group. This difference was statistically significant (P = 0.016).

**Figure 2.** Comparison of Global Symptomatic Score (GSS) in the intervention and control groups at the beginning of admission, days 3, 7 and 14 after admission.

### Evaluation of blood oxygen saturation of patients in two groups

There was no significant difference in blood oxygen saturation between the two groups at the beginning of the study (92.07 ± 4.6 in the control group vs 91.56 ± 3.39 in the intervention group). As can be seen in Figure 3, blood oxygenation improved in both groups and the slope of oxygenation was higher in the ethanol group. Although, this difference is not statistically significant (P = 0.097).

**Figure 3.** Comparison of mean blood oxygen saturation (SPO2) in intervention and control groups at the beginning of admission, days 3, 7 and 14 after patient admission.

### The effect of intervention on inflammatory factor (CRP)

CRP had a decreasing trend in both groups, significant difference was not seen (P = 0.276).

**Figure 4.** Comparison of CRP in the intervention and control groups at the beginning of admission and three days after patient admission.

### Clinical Status

Clinical status based on the modified 7-point ordinal scale was compared in two groups on day 14 of the study, the results of which were based on Table 3.

**Table3:** Comparison of clinical status scale (CSS) of intervention and control groups on the 14th day of admission.

| **Control N=55** | **Intervention N=44** | **Characteristic and Score N (%)** |
| --- | --- | --- |
| 0 | 0 | 1. Death |
| 0 | 0 | 2. Hospitalized, on invasive mechanical ventilation |
| 0 | 0 | 3. Hospitalized, on non-invasive ventilation or high flow oxygen devices |
| 2 (3.63) | 0 (0) | 4. Hospitalizations for any reason and need oxygen |
| 10 (18.18) | 2 (4.54) | 5. Requiring ongoing medical care or supplemental oxygen at home |
| 29 (52.72) | 13 (29.54) | 6. Continue signs or symptoms without requiring supplemental oxygen - no longer requires ongoing medical care |
| (25.45) | 29 (65.90) | 7. Complete recovery |

**Figure 5.** Comparison of Clinical Status on a 7-Point Ordinal Scale (CSS) on Study Days 14 in two groups

The odds of intervention group have different clinical status was 5.715 (95% CI, 2.47 to 13.19) times that of standard group, a statistically significant effect, Wald χ2 (1) = 16.67, p <.001. After end of the treatment period in the control group, 6 patients (10.9%) were readmitted to receive additional treatment or hospitalization. In the ethanol group, none of the patients was readmitted, P=0.02.

### Adverse events and safety

Six out of 50 patients in the ethanol group (12%) interrupted the treatment because of adverse events at the onset of inhalation and, thus, were excluded from the statistical analysis No more than one case was observed for every side effect and that side effect disappeared after stopping ethanol use. Adverse events included: hiccups, eye irritation, cough, shortness of breath, sneezing and unpleasant odor of alcohol.

## Discussion

The idea of inhalational ethanol therapy has been proposed based on its viricidal properties causing dissolution of the virus fat layer and inhibiting its proliferation, in addition to mitigating hyperactivity of the immunity system for COVID-19. Based on this information, several studies have been conducted on this topic. Patients with positive RT-PCR test, moderate clinical symptoms, and indicated for Remdesivir treatment according to the Ministry of Health of Iran protocol, were enrolled in this study. The results show that in the intervention group (ethanol therapy) the clinical symptoms decreased more than in the control group and more rapidly, these data reached the statistical significative level

Regarding others treatment outcomes, the response of the ethanol group was better because no patient was readmitted but in the control group (placebo, whereas 6 patients (10.8%) in the control group needed to repeat the standard treatment or needed to be hospitalized. Moreover, the exaggerated immune system response was further inhibited by ethanol therapy and the rate of reduction of inflammatory factor CRP was higher in this group (P = 0.05). In the ethanol group, blood oxygenation improved faster and the slope of oxygen increase was higher in the ethanol group, although there was no statistically significant difference between the two groups (P = 0.097). These findings may support that ethanol is virucidal. The faster and more rapid reduction of CRP in this study shows that the hypothesis of inhibition of the immune system with ethanol is correct. The purpose of prescribing expensive medications such as tocilizumab or steroids is also to inhibit immunity.

## Conclusion

Looking at the efficacy of the inhaled nebulized ethanol, its use seems to be effective in general rapid improvement, mitigating clinical symptoms and reducing the need to repeat treatment. Considering the low cost, availability and no significant adverse events of ethanol, research and additional efforts are recommended to evaluate its curative and preventive effects in the early stages of COVID-19.

## Data Availability

All data produced in the present study are available upon reasonable request to the authors

## Acknowledgments

Our thanks go to consultation and scientific relation with other countries scientist, especially Prof. Tsumoru Shintake (Okinawa Institute of Science & Technology Graduate University, Okinawa, Japan), Thomas Manning (Ph.D. Professor of Chemistry in Valdosta University), Dr. Steven Stogner (Pulmonologist, Forrest General Hospital, Hattiesburg, MS, USA), Dr. Ali Shahshahan (Internist, Hattiesburg Clinic, Mississipi) and Annemarie Padmos (Master of Advanced Nursing Practice, Netherlands). We are also grateful to coworking Dr. MR Nazer (Infection Disease Specialist) and Dr. S. Abbasi (Intensivist) and other Nurses in ethanol therapy project in Isfahan.

